# Unusually high frequency of dengue cases with warning signs in patients with no history of previous dengue infection at the Santa Rosa Hospital in the city of Lima

**DOI:** 10.1101/2025.01.30.25320345

**Authors:** Solomon Durand, Nadia Wong, Estefanía Briceño, Raquel Delgado, Yolanda Sánchez, Maria Huamani, Judith Fabian, Jean Flores, Pedro Contreras

**Affiliations:** Santa Rosa Hospital; Universidad Nacional Mayor de San Marcos

**Author notes:** **Corresponding author** Salomón Durand, mail. **Funding Source** Self Financed.

**Keywords:** Severe dengue, Dengue Virus Pathogenicity, Dengue complications, Dengue classification

## Abstract

**Introduction:** **In** 2024, a dengue epidemic affected the city of Lima, a city of more than 10 million inhabitants, for the first time. The Santa Rosa hospital treated more than 5000 febrile patients between January and July 2024, identifying 1788 patients with dengue, 18.6% of them with warning signs or serious, despite being the first infection for them. In order to know the factors related to this unusually high frequency of cases with warning signs and severity in a group of patients who were infected for the first time, this study was developed.

**Methodology:** The epidemiological records were reviewed and contrasted with the information from the hospital’s emergency care records. The laboratory results were obtained from the laboratories of the hospital and reference in Lima.

**Results:** Between January and July 2024, 5,862 febrile patients presented to the hospital emergency room, 1788 were considered cases of dengue. Nine cases (0.5%) were classified as severe dengue and 324 (18.1%) as dengue with warning signs. 4 died. 9.3% of patients over 65 years of age presented some warning or severity sign, while 6.4% of patients under 65 years of age presented warning signs (P 0.05). Only 2.5% (45/1788) responded that they had previously had dengue, but there was no difference between those who presented warning signs or severity.

**Conclusions:** In conclusion, the dengue epidemic in Lima reached a greater severity than expected despite being the first infection for the affected population, a new massive outbreak and the entry of a new serotype could further increase the severity of the cases.

## Introduction

In Peru, there have been reports of transmission of the dengue virus (DENV) since 1990, behaving as an endemic with an increase in cases in the rainy season or as an outbreak or epidemic upon the entry of a new serotype or genotype, such as the one that occurred in 2011, when the Asian/American genotype of DENV II was introduced in the city of Iquitos. which then spread throughout the Amazon and northern coast of Peru, presenting great morbidity and lethality ^(1) (2,3)^

Unlike the rest of the country’s regions, the city of Lima had not been affected by a dengue epidemic until 2024. Although it is true that in 2000, the reintroduction of Aedes aegypti had already been reported in Lima and the first outbreak of dengue was registered in 2005, these outbreaks were isolated and self-limiting with the onset of winter in Lima. However, in 2024, the city of Lima was affected by an unprecedented outbreak of dengue, surpassing Lima for the first time in number of cases (87,511 and 261,268 in Lima and Peru respectively) than other regions of Peru. In 2024, 40 of the 43 districts of Lima had dengue transmission, and an outbreak had never affected almost the entire city, even when the major dengue epidemics occurred in Peru in 2011 and 2017 (Figure 1). ^(4) (5) (6)^

During 2023, 274,246 cases of dengue and 442 deaths were reported in Peru, in this same period Lima reported 27,574 cases. From 2024 to September, 261,268 cases of dengue have been reported in the country and this year Lima has almost a third of the cases reported in the entire country ^(6).^

In this context, it could be stated that dengue in Lima was presented as an epidemic recently and therefore for the population it would be the first infection, so we would expect a lower severity of the clinical picture, considering that it is the second infection that carries a risk of severity, a situation different from that of the Amazon and the northern coast of the country where probably most of the population has already had contact with one or more serotypes of DENV due to transmission and outbreaks recorded in these regions for 20 years ^(7,8)^

The Santa Rosa hospital is located in a district of Lima of medium or high socioeconomic status, with homes that have drinking water services and modern infrastructure, however, during 2024, an increase in dengue cases was observed in the hospital by 8 times compared to 2023, 230 in 2023 and 1788 until July 2024. ^(9)^

At the Santa Rosa Hospital, in 2024, due to the increase in febrile patients seeking care in the hospital’s emergency, a tent was set up for the care of febrile patients where dengue was ruled out and an intensive surveillance unit for dengue cases (UVICLIN) was also organized with 15 beds. To facilitate the registration and notification of cases, an electronic epidemiological file was developed that was filled out by the same doctor who provided care.

According to information from the hospital’s epidemiology office, 18.6% of the dengue cases that attended the hospital’s emergency room had warning signs or severity at the initial consultation, which is not expected in a DENV epidemic that affects a population for the first time.

For this reason, it was decided to carry out a study with the aim of knowing the factors related to the unusually high frequency of dengue cases with warning signs treated at the Santa Rosa hospital during 2024 and also to analyze and document the change in the demand for dengue in the Hospital and in the cases in the city of Lima.

## Methodology

The study was carried out at the Santa Rosa Hospital, which is located in the district of Pueblo Libre located in the center of Lima. The city of Lima is the largest and most populous in Peru, it has 10,200,000 inhabitants, and is divided into 43 districts, 7 of them are in the area of the Santa Rosa hospital, these districts are characterized by being of high socioeconomic strata. ^(9)^

The study had an analytical, descriptive and retrospective design. For the purpose of the study, the electronic epidemiological file was reviewed together with the manual epidemiological files that existed, with which an Excel database was built. The information in this database was completed and contrasted with the information in the electronic emergency care records. Likewise, the database of laboratory results of dengue discard tests of the hospital and the reference laboratory of the Directorate of Integrated Health Networks of Central Lima were reviewed.

Patients with a positive result to a rapid test (positive IgM or NS1) or to an ELISA for NS1 or IgM or positive PCR were considered as cases, however, those patients with compatible clinical symptoms and with an epidemiological link were also assumed as cases.

Patients were classified according to severity as no warning signs, with warning signs or severe according to the World Health Organization’s dengue case care guideline ^(10)^

To prepare the table of signs and symptoms on admission, only the electronic epidemiological record was considered because it is a first-hand source. The database was anonymized prior to statistical analysis.

To determine the number of dengue cases in Lima, the weekly epidemiological bulletins of the Center for Disease Prevention, available online since 1998, were reviewed.

The data were summarized with measures of central tendency and the risks of severe dengue were analyzed with a univariate analysis, calculating the OR and its confidence limits using SPSS software.

The study was approved by the research ethics committee of the Santa Rosa Hospital with the certificate of approval 043-2024-CEI-HSR.

## Results

Between January and July 2024, 5,862 fevers occurred in the emergency room of the Santa Rosa hospital, and 1893 cases were considered probable cases of dengue, of them, 1788 cases of dengue were confirmed, 778 (43.5%) by a laboratory test and the rest by the clinical picture and epidemiological link, 105 cases were discarded.

Nine cases (0.5%) were classified as severe dengue in the emergency and 324 (18.1%) as dengue with warning signs. For the analysis, cases with severe dengue and warning signs were grouped into a single group, 333 cases with dengue with warning signs and severe (18.6%). During this period, 4 people with severe dengue died, all of them over 65 years of age.

9.3% of patients over 65 years of age presented some warning or serious sign, while 6.4% of patients under 65 years of age presented warning signs (P 0.05), (Table No. 1)

**Table 1.** Main demographic variables of dengue patients admitted to Santa Rosa Hospital in 2024.

|  | Dengue s/warning signs | Dengue w/warning signs | Grand total | OR | 95% CI | P-Value |
| --- | --- | --- | --- | --- | --- | --- |
|  | n(%) | n(%) | N(%) |  |  |  |
|  | 1455 (81.4) | 333 (18.6) | 1788 (100) |  |  |  |
| Age (years)* | 29[21-45] | 27[19-45] | 29 [21-45] |  |  |  |
| <=65 years | 1362 (93.6) | 302 (90.7) | 1664 (93.1) | 1 |  |  |
| >65 years old | 93 (6.4) | 31 (9.3) | 124 (6.9) | 1.503 | [0.983-2.300] | 0.059 |
| Sex |  |  |  |  |  |  |
| Female | 891 (61.2) | 193 (58.0) | 1084 (60.6) | 1 |  |  |
| Male | 564 (38.8) | 140 (42.0) | 704 (39.4) | 1.146 | [0.900-1.459] | 0.269 |
| Place of origin |  |  |  |  |  |  |
| Districts of the jurisdiction | 860 (59.1) | 184 (55.3) | 1044 (58.4) | 1 |  |  |
| Other districts | 595 (40.9) | 149 (42.9) | 744 (41.6) | 1.170 | [0.921-1.488] | 0.199 |
| Had dengue in the past |  |  |  |  |  |  |
| Yes | 35 (2.4) | 10 (3.0) | 45 (2.5) | 1.339 | [0.650-2.758] | 0.428 |
| No | 778 (53.5) | 166 (49.8) | 944 (52.8) | 1 |  |  |
| S/D | 642 (44.1) | 157 (47.1) | 799 (44.7) |  |  |  |
| Co-morbidities |  |  |  |  |  |  |
| Yes | 129 (8.9) | 31 (9.3) | 160 (8.9) | 1.124 | [0.730-1.731] | 0.597 |
| No | 664 (45.6) | 142 (42.6) | 806 (45.1) | 1 |  |  |
| S/D | 662 (45.5) | 160 (48) | 822 (46.0) |  |  |  |

\*Median [interquartile range]
Source: Dengue Database – HSR. 2024

Table No. 2 shows the signs and symptoms found in the first care. Abdominal pain was the most frequent warning sign (20.3%), followed by vomiting (7.4%) and mucosal bleeding (3.6%).

**Table 2.**
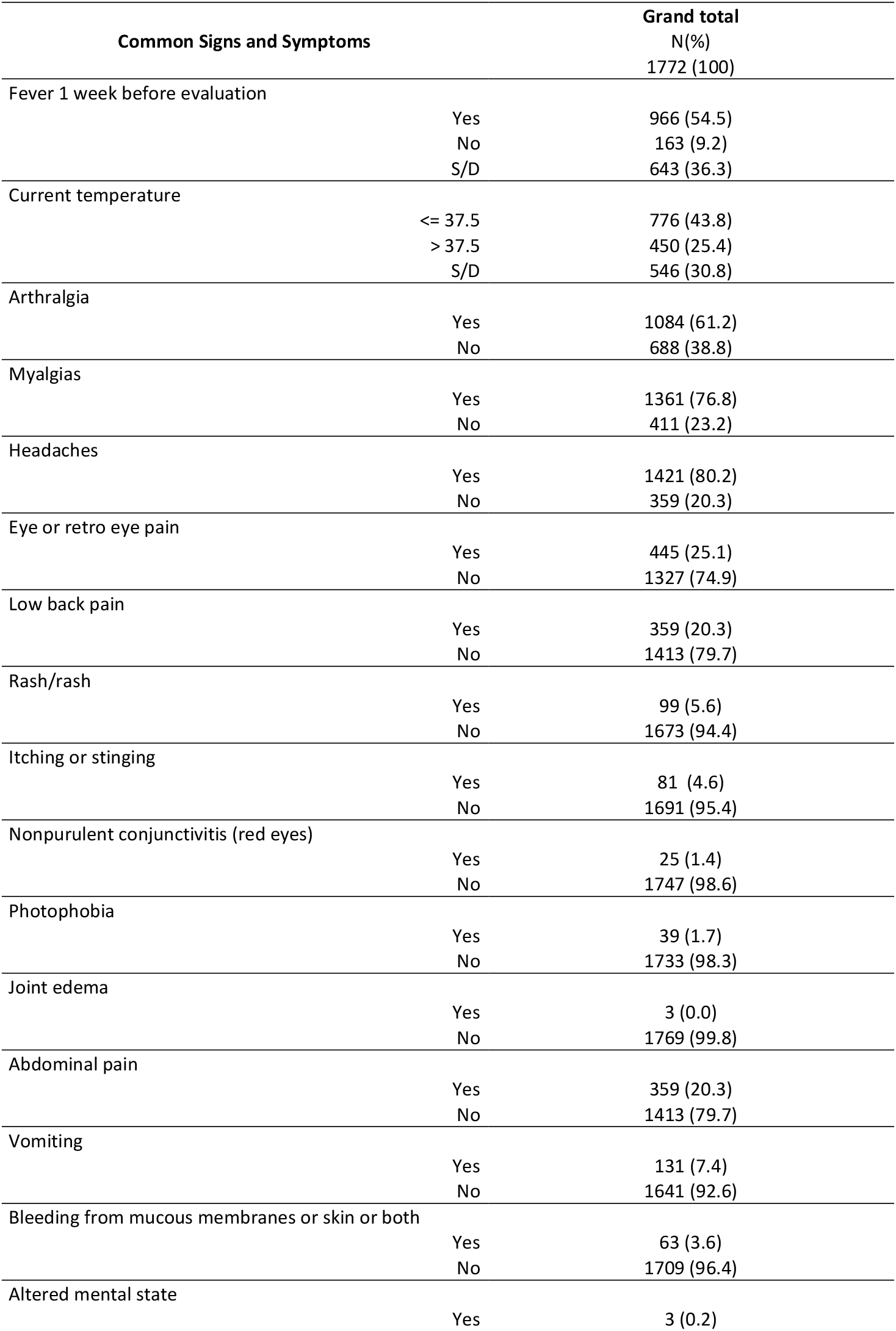
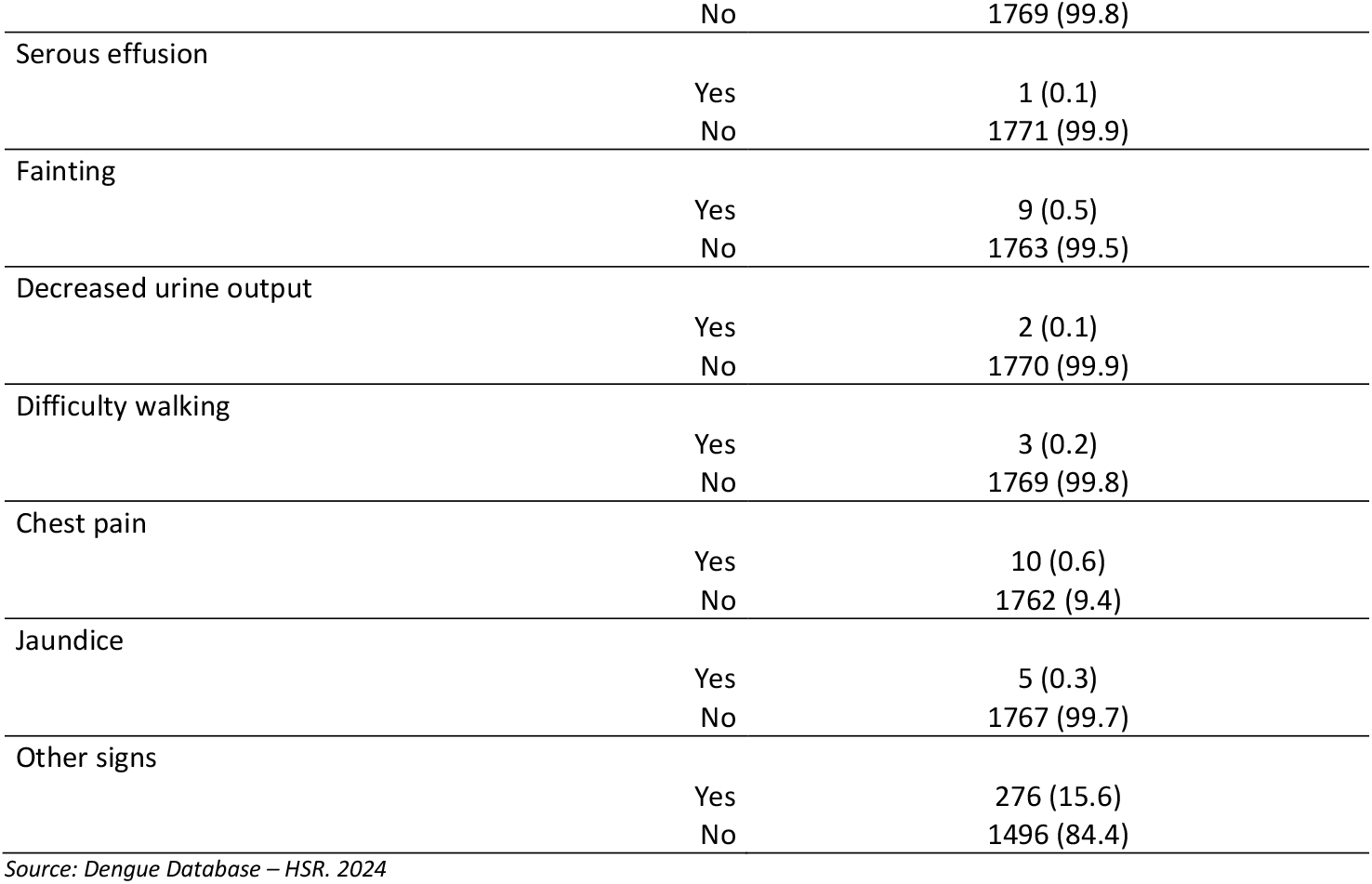
Frequent signs and symptoms of dengue patients admitted to Santa Rosa Hospital in 2024.

**Table 3.**
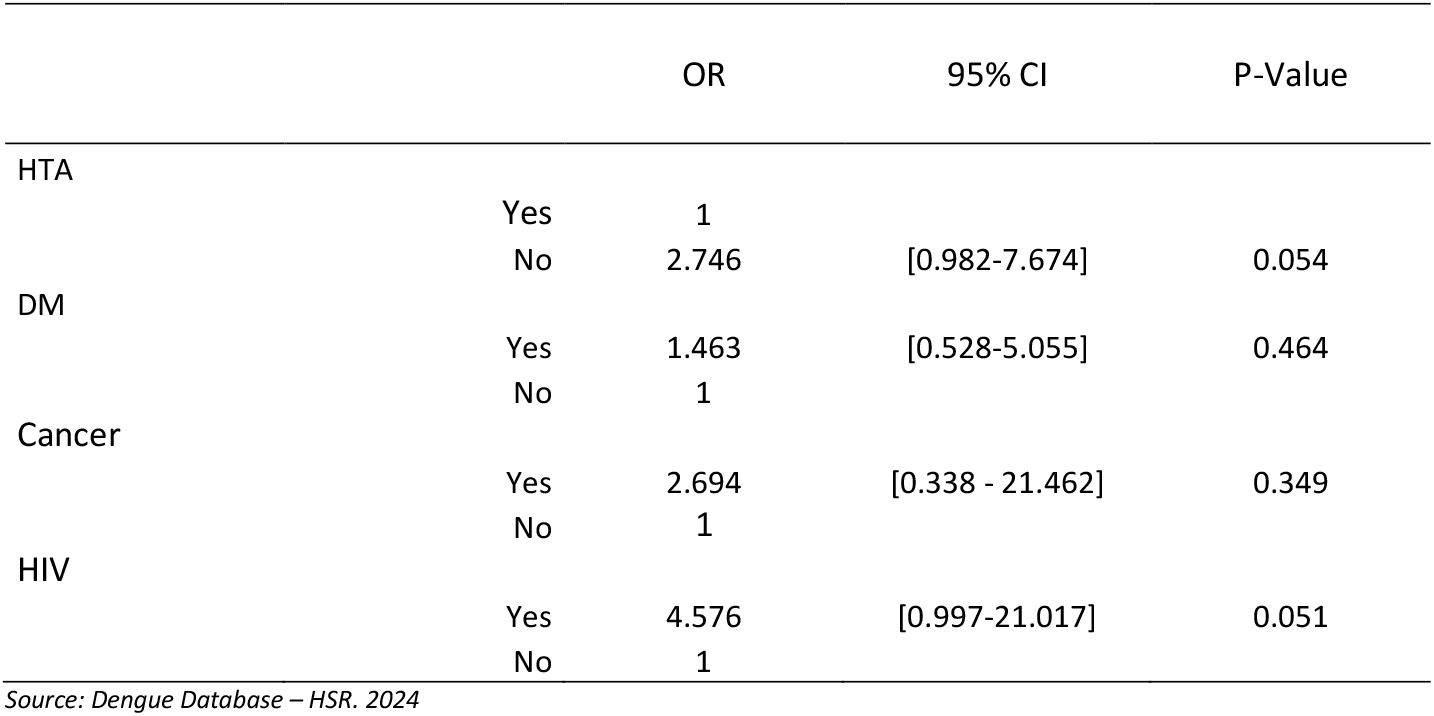
Main co-morbidities of dengue patients admitted to the Santa Rosa Hospital in 2024.

|  |  | OR | 95% CI | P-Value |
| --- | --- | --- | --- | --- |
| HTA | Yes | 1 |  |  |
|  | No | 2.746 | [0.982-7.674] | 0.054 |
| DM | Yes | 1.463 | [0.528-5.055] | 0.464 |
|  | No | 1 |  |  |
| Cancer | Yes | 2.694 | [0.338 - 21.462] | 0.349 |
|  | No | 1 |  |  |
| HIV | Yes | 4.576 | [0.997-21.017] | 0.051 |
|  | No | 1 |  |  |

Source: Dengue Database – HSR. 2024

58% of the patients with warning signs were female, but there was no difference with respect to sex with the group without warning signs

Only 2.5% (45/1788) responded that they had had dengue before, there was no difference between those who presented warning signs or severity 3% and those who did not present 2.4%.

A total of 68 patients (3.8%) were positive for IgG, 14 in the group with warning signs and 1 in the severe group (4.2% in the group with warning signs and severe) and 53 (3.6%) in the group without warning signs.

Regarding comorbidities, 8.9% were found to have some comorbidity, the most frequent being diabetes mellitus and arterial hypertension, no difference was found in the univariate analysis of comorbidity with respect to higher risk of dengue but rather lower risk in the case of hypertension.

## Discussion

In this study, it was found that the dengue epidemic in the city of Lima, which occurred in 2024, had an unusual frequency of cases with warning signs (18.6%), and age over 65 years was the main risk factor related to the presentation of warning signs or severity.

The epidemic transmission of dengue in Lima is recent. Unlike the Amazon and northern coast where the DENV has been circulating since 1990, in Lima there had been isolated and self-limiting outbreaks. In the year 2000, the presence of Aedes aegypti was reported in Lima and five years later, in 2005, an outbreak occurred in North Lima that was possibly controlled by the limited dispersion of the vector and the low temperatures in Lima in the winter months. The neighborhoods where the outbreaks occurred were characterized by a lack of water in the home, which forced water to be collected in containers. The entry of Asian/American serotype 2 in 2010 produced an epidemic in several regions of Peru, but it did not affect the city of Lima nor was it affected in 2017, when there was a major outbreak on the north coast (graph No. 1). However, the Aedes was gradually dispersing in the city, there is currently a presence of the Aedes in 41 of the 43 districts that make up Metropolitan Lima, the dispersion of the vector created a suitable scenario for a dengue epidemic to begin in Lima in 2023 that covered almost the entire city. (11)

**Graph N° 1.**
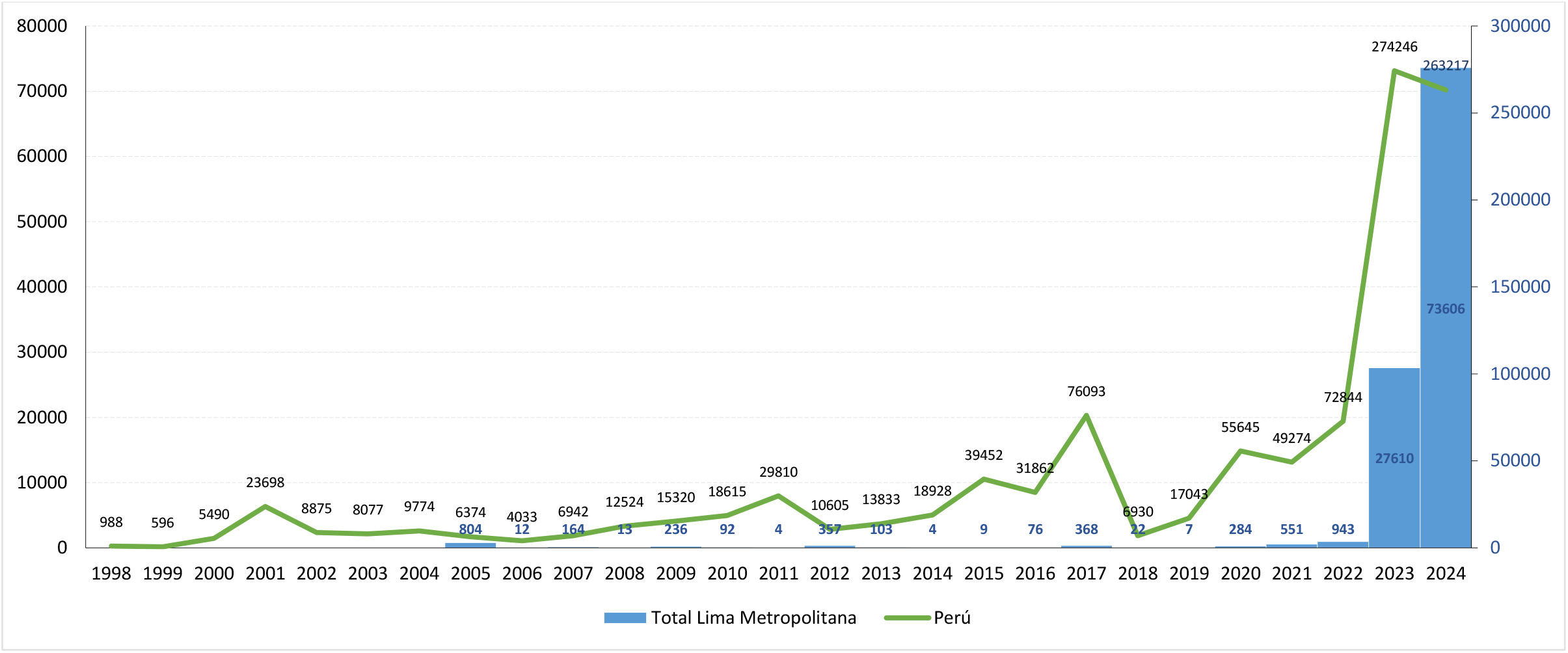
Dengue in Peru and Lima (1998-2024) *Source: CDC Epidemiological Bulletins – MINSA Peru. * until week 38 of 2024*.

The few cases of severe dengue identified (9.0.5%) are striking, but it should be taken into account that the epidemiological files are filled out at the first contact with the patient and it is possible that the signs of severity may have occurred during the evolution, according to Durand et al. in an ICULCIN of the Iquitos Hospital, up to 30% of the cases admitted with warning signs evolved to severity during hospitalization. ^(12)^

Dengue transmission in Lima is recent, and based on the CDC epidemiological bulletins available at WEB small and sporadic outbreaks have been observed in Lima since 2005 (Figure 1). At the Santa Rosa Hospital, before 2023, very few cases were treated, 5 in 2020, 8 in 2021 and 13 in 2022, which went to 1788 in 2024.

Only 2.5% of patients had a history of dengue in the epidemiological file and no differences were found between exposure to the virus and the risk of having dengue with warning signs or severe, although the sample of those who had a history of dengue is too small to find the differences. If we take IgG positives as cases with a history of previous dengue, the number is also not important 3.8% (68/1788), 14 positive IgG in the group with warning signs and 1 severe (4.2%) and 53 (3.6%) positive IgG in the group without warning signs.

However, the finding is important, because this result would make us look for another factor other than the previous infection to explain the unusually high percentage of dengue with warning signs and severe in the epidemic in Lima. The importance of the history of dengue as a factor of severity was proven in epidemiological studies carried out since the 90’s in Southeast Asian countries where it was found that children who had had dengue had more frequent severe dengue, later these findings were corroborated in epidemiological studies carried out in Cuba and Nicaragua. The pathophysiology of the higher frequency of severity in the second infection is not well understood, but it is postulated that a higher and longer viremia occurs in the second infection due to the binding of non-neutralizing antibodies to the virus, followed by their uptake in monocytes carrying the Fc receptor (antibody-dependent increase), an exacerbated immune response could also occur, by inflammatory mediators that contribute to intravascular leakage, triggered by activated natural killer cells and memory T cells. The nonstructural protein NS1 secreted by infected cells is associated with vascular leakage by damaging the endothelial glycocalyx and altering endothelial cell junctions. This process could be exacerbated during secondary infection due to increased viremia. (7,8,13) However, as we see in this study, the virulence of the strain also plays an important role in the clinical course of the disease.

On the other hand, it was not found in this study that the presence of any comorbidity increased the possibility of having dengue with warning signs, however, age is a risk, people over 65 years of age have a higher risk (OR 1.5) in the univariate analysis.

Genotype probably played an important role in the severity of the symptoms regardless of previous infection. In the 2011 epidemic, when the American/Asian DENV 2 was admitted, the percentage of patients requiring hospitalization increased in the Iquitos hospital (18.2 with serotype 2 and 9.2% with serotype 4), and this would be related to the DENV genotype. In Lima in 2024, until June, 69.5% of dengue cases were due to serotype 1, 30% to serotype 2 and 0.4% to serotype 3. (12) (14)

According to the literature, the factors that would be related to severity or death from dengue are: the virulence of the infecting strain (serotype, genotype and viral load), the immunity of the host (previous infection, sequence of infecting serotypes, previous serotype), the factors of the host (comorbidity, age, gestation) and the quality of care received and the opportunity to seek medical attention by the patient would also be factors. In this study, the virulence of the strain, the age of the host and the opportunity to seek attention would be factors. (8)

The outbreaks that occurred in Lima before 2023 were localized and self-limiting, however in the latter transmission occurred in almost all of Lima, this is not well explained, because the previous outbreaks occurred in communities that had poor water distribution and where water accumulated in containers, but it is not explained how in a region where it never rains and in districts where there is adequate water distribution and relatively modern housing, has become accustomed to the vector. It is necessary to study which would be the habitats that enable the permanence of the vector in mesocratic districts such as pueblo libre or Jesús María. ^(9)^

The limitations of this study would be that it was not possible to evaluate the patient’s evolution, probably many of them developed signs of severity and information on laboratory results such as platelet count, etc., was not recorded. They were not available in databases. Even so, what has been found is important to prevent severity and death in the future.

In conclusion, the dengue epidemic in Lima reached a greater severity than expected despite being the first infection for the affected population, a new massive outbreak and the entry of a new serotype could further increase the severity of the cases.

## Data Availability

All data produced in the present study are available upon reasonable request from the authors.

